# Clinical and Demographic Features of Distal Extremity Weakness in Myasthenia Gravis - A Scoping Review

**DOI:** 10.1101/2024.08.16.24312046

**Authors:** Emrah Kacar, Mahavishnu Sahadevan, Maria Isabel Leite

## Abstract

**Introduction:** Myasthenia gravis (MG) is a chronic autoimmune neuromuscular disease primarily affecting extraocular, bulbar, axial, and proximal extremity muscle groups. However, a subgroup of patients experience distal extremity weakness, known as distal myasthenia gravis (DMG). Despite its longtime observation, DMG’s clinical and demographic features remain poorly understood. This scoping review aims to summarise current evidence regarding the demographic, clinical, and diagnostic features of autoimmune MG patients who exhibit atypical, distal extremity weakness.

**Methods:** We conducted a scoping review by searching the following electronic databases: Embase, Medline (Ovid), Scopus, Web of Science, and ResearchGate, covering the period from their inception up to August 2024. We included all types of publications due to the limited number of articles available on DMG. Since we wanted to work on individual-level patient data, we selected papers based on our inclusion and exclusion criteria, and interpreted the data descriptively.

**Results:** After the screening of the initial 2162 publications, we assessed 55 for eligibility. 23 articles (11 case reports, 5 letters to the editor, 2 poster abstracts, and 5 case series) fulfilled the inclusion and exclusion criteria. Demographic and clinical details of 43 patients were extracted for further analysis. The male-to-female ratio was 1.4:1, with a mean (SD) age of MG onset at 40.2 ± 17.2 and a mean age of DMG onset at 44.9 ± 16.3. As 38% DMG patients developed distal extremity weakness at disease onset, in total 55% of DMG patients showed distal symptoms within the first year of their diagnosis. In women with DMG, generalised MG was often present at disease onset (83%), whereas men had similar percentages of ocular (52%) and generalised (48%) MG at disease onset. The most frequently reported weaknesses were in finger extension, followed by wrist extension, elbow extension, and finger abduction. Ankle dorsiflexion weakness was the most frequently reported form of distal lower extremity weakness.

**Conclusion:** Distal myasthenia gravis (DMG) is a rare yet debilitating subtype of myasthenia gravis. When evaluating MG patients with distal symptoms, it is crucial to consider various differential diagnoses resulting in distal extremity weakness. In this context, we would like to emphasise two MG-associated conditions, namely myasthenia-inflammatory myopathy (MG-IM) and myasthenic neuromyopathy. Our scoping review highlights the need for future prospective studies to address unanswered questions to improve clinical outcomes and the quality of life for patients with DMG.

**Highlights:**

- Significant weakness of distal extremity muscles is rare (3-7%) in myasthenia gravis (MG) and is known as distal myasthenia gravis (DMG).
- Finger and wrist extensors are the most frequently reported distal muscles affected in DMG.
- Approximately 38% of the DMG patients in this review experienced distal extremity weakness at the onset of MG, and a total of 55% developed distal symptoms within the first year of the disease.
- At disease onset, 83% of women with DMG presented with generalised MG, whereas men had similar percentages of ocular (52%) and generalised onset (48%).
- When evaluating MG patients with distal symptoms, it is crucial to clinically consider two MG-associated conditions, namely myasthenia-inflammatory myopathy (MG-IM) and myasthenic neuromyopathy.

## 1. Introduction

Myasthenia gravis (MG) is a chronic autoimmune disorder in which autoantibodies impair the communication between nerves and muscles, resulting in weakness and fatigability of the latter [1]. A systematic review reported a crude estimated pooled incidence rate of 5.3 cases per million person-years and a prevalence rate of 77.7 cases per million, with a population-dependent variation observed globally [2]. MG affects individuals of any age and its incidence increases with age in both sexes with seemingly male predominance in the elderly [2,3]. Therefore, understanding diverse phenotypes in MG is more important than ever in our ageing population.

People with MG (pwMG) most commonly present with extraocular, bulbar, axial, and proximal limb muscle weakness [4]. Dysfunction of distal extremity parts, such as muscles affecting hand and foot movements, has occasionally been reported in the literature over the years. Even early clinical descriptions of myasthenia gravis (MG) noted weakness in the hands and forearms among the probable initial symptoms [5]. Two retrospective studies on DMG found that 3% (9 of 236) and 7% (6 of 84) of patients with myasthenia gravis in their respective cohorts exhibited distal extremity weakness [6,7]. The muscles most affected in both studies were the finger extensors, with additional involvement of the wrist extensors, finger abductors, wrist flexors, elbow extensors, elbow flexors, and ankle dorsiflexors.

Previous literature on distal myasthenia gravis (DMG) have focused primarily on the weakness of the muscles of the hands and feet [8]. In our primary literature search, we encountered case series involving myasthenia gravis (MG) patients of African-American, Asian, and Indian ancestries, which showed less common biceps and triceps muscle weakness [9–11]. Since these cases did not align with the typical MG phenotype, we expanded the definition of distal weakness in our scoping review to include biceps and triceps muscle weakness.

Distal limb weakness significantly impacts the quality of life of patients with MG [12]. However, clinical and demographic characteristics of DMG patients have not been thoroughly investigated, and current clinical MG scores do not provide a detailed assessment of dysfunction in distal muscle groups [13]. For this reason, the objective of our article is to review the existing literature on demographic, clinical and diagnostic features of autoimmune MG patients with atypical, distal extremity weakness, who are not experiencing a myasthenic crisis.

## 2. Methods

We conducted a comprehensive literature search in August 2022, updated it in February 2023 and August 2024, with the most recent search conducted on August 8, 2024. We used several electronic bibliographic databases, including Medline (Ovid), Embase, Scopus, Web of Science and ResearchGate. Additionally, we performed citation searching based on the references of articles of interest. An overview of our search process and the reasons for exclusions are listed in Fig. 1. Since all analyses are based on using previously published studies, ethical approval and patient consent are not required.

**Fig. 1:**
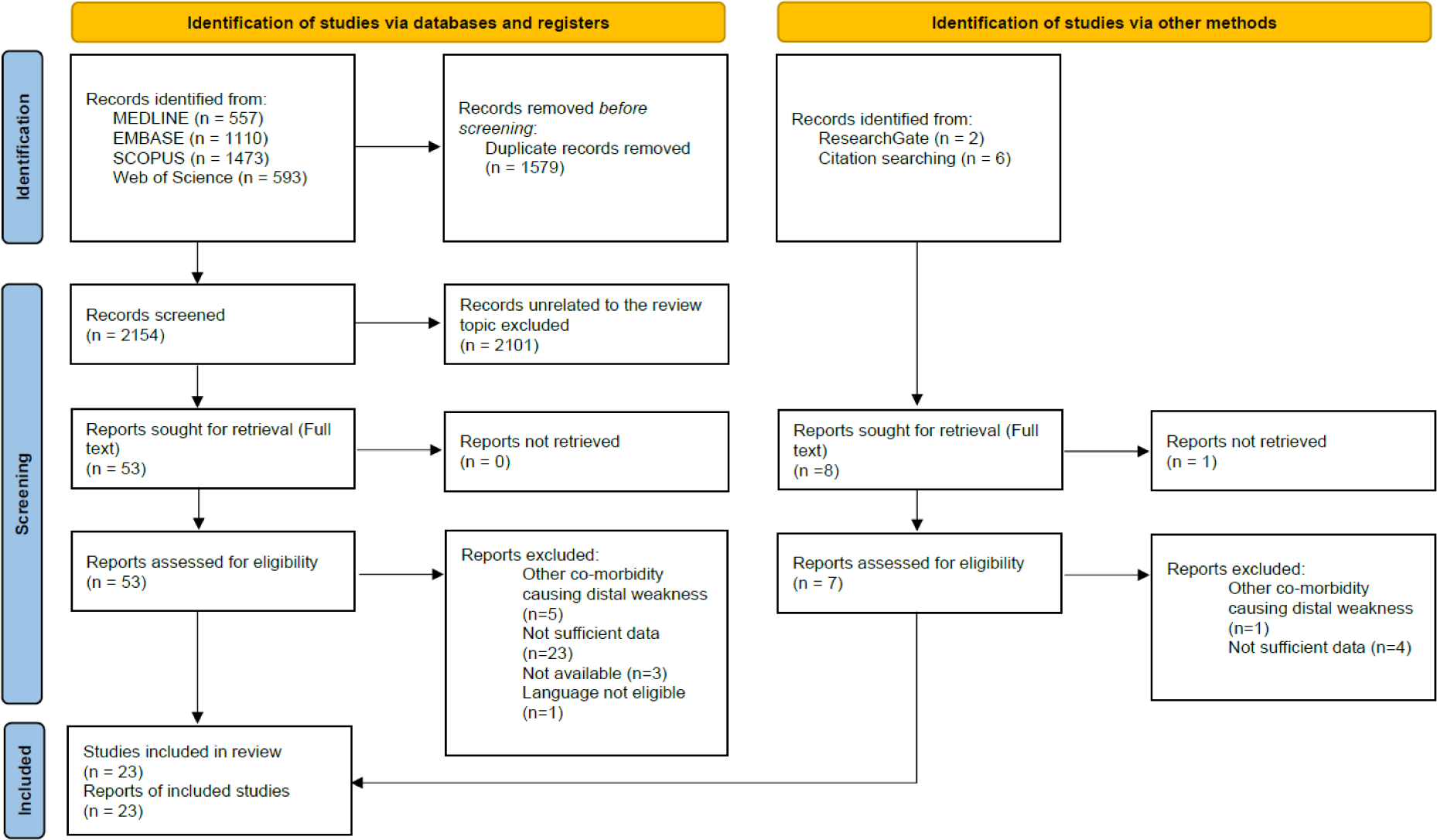
PRISMA flow diagram.

### 2.1. Search strategy

Our search strategy followed a three-step method as recommended in standard JBI scoping reviews [14]. First, we conducted an initial limited search in Medline (Ovid) and Embase. Then, we analysed the common text words contained in the title and abstracts as the foundation for our search strategy. Subsequently, we undertook a second search using that foundation in all included databases. Thirdly, we searched the reference list of all identified reports and articles for additional papers. Publications in English and Portuguese were included. Search strategy was peer-reviewed by an Oxford Bodleian librarian using the Peer Review of Electronic Search Strategies (PRESS) checklist. The search strategy can be found for different databases in the Appendix under “Search Strategy”.

### 2.2. Inclusion and Exclusion Criteria

We included studies of autoimmune MG patients across all age groups, given they met the following criteria: (1) included individual data on demographic and clinical features, electrophysiology studies, antibody status, (2) fulfilled the clinical criteria for MG [15] and (3) had clinically confirmed atypical, distal extremity weakness resulting from MG. Distal extremity weakness included muscle groups responsible for elbow extension and flexion; wrist extension and flexion; finger extension, flexion, abduction, and adduction; thumb abduction, adduction, flexion, extension and opposition; ankle and toe dorsiflexion and ankle plantar flexion. Studies that included patients with congenital myasthenia or distal extremity weakness associated with Lambert-Eaton syndrome, Guillain-Barre Syndrome, polyneuropathy, myositis, muscle dystrophies, traumatic injury, rheumatoid arthritis, and motor neuron disease, as well as those containing patients in a myasthenic crisis, were excluded.

### 2.3. Article Selection

Two neurologists (E.K. and M.S.) independently screened all searched articles’ titles and abstracts. The deduplication and the screening for inclusion and exclusion was conducted with Rayyan Software[16]. Both reviewers independently reviewed the full text of eligible articles and any disagreement on inclusion or exclusion were resolved through discussion. Out of the 55 reports assessed in full-text for eligibility, 23 were excluded due to insufficient clinical data [7,8,17–37], 5 due to other possible causes for distal weakness [38–42], 3 due to unavailability [19,43,44] and one due to language not eligible for inclusion [45]. In summary, 23 reports met the inclusion and exclusion criteria and were considered for further analysis. A list of these reports and their web links can be found in Supplementary.

### 2.4. Data Extraction

One neurologist (E.K.) conducted the data extraction, which was then verified by the other neurologist (M.S.). The data extracted consisted of various components, including the year of publication, first author, publication type, age at onset of myasthenia gravis (MG), age at onset of distal myasthenia gravis, sex, acetylcholine receptor (AChR) antibody status (positive/negative), symptoms at MG onset (ocular or generalised), time from MG onset to first distal weakness (in weeks, months, or years), electromyography (EMG) results (normal or abnormal (myopathic changes)), repetitive nerve study (positive or negative or not reported), single fiber-EMG (positive or negative or not reported), predominant weakness (upper or lower extremity), treatment types and treatment response. Demographic and clinical characteristics of included DMG patients are shown in Table 1.

**Table 1:** Demographic and clinical characteristics of selected DMG patients.

| Pat. No. | Year | Ref. | Pub. | Age (MG-Onset) | Age (DMG-Onset) | Sex | AcR-A b | MG-Type at Onset | Time to DMG | EMG | RNS | SFEMG | Pred. W | Tx | Tx Resp. | OCEBM |
| --- | --- | --- | --- | --- | --- | --- | --- | --- | --- | --- | --- | --- | --- | --- | --- | --- |
| 1 | 1997 | Uncini et al.[55] | LTE | 69 | 69 | F | + | Generalised | 0 | Normal | + | NR | LE | AChEI, PDN | Yes | 5 |
| 2 | 1998 | Janssen et al.[46] | CSN | 76 | 76 | M | - | Ocular | 6 m | Normal | + | + | UE | AChEI, PDN | Yes | 5 |
| 3 | 1999 | Nations et al.[6] | BC | 21 | 33 | F | + | Ocular | 12 y | NR | + | NR | UE | NR | Lost to FU | 5 |
| 4 |  |  |  | 33 | 33 | F | + | Generalised | 0 | NR | + | NR | UE | NR | Yes |  |
| 5 |  |  |  | 35 | 45 | F | + | Ocular | 10 y | NR | + | + | UE | NR | Yes |  |
| 6 |  |  |  | 25 | 25 | F | + | Generalised | 0 | NR | NR | + | UE | NR | Yes |  |
| 7 |  |  |  | 30 | 33 | M | + | Generalised | 3 y | NR | + | NR | UE | NR | Lost to FU |  |
| 8 |  |  |  | 25 | 25 | F | + | Generalised | 0 | Normal | + | NR | UE | NR | Yes |  |
| 9 |  |  |  | 14 | 59 | M | + | Ocular | 45 y | Normal | + | NR | UE | NR | Yes |  |
| 10 |  |  |  | 36 | 36 | M | + | Ocular | 1 m | NR | + | NR | UE | NR | Yes |  |
| 11 |  |  |  | 29 | 43 | M | + | Generalised | 14 y | Normal | + | + | UE | NR | NR |  |
| 12 | 2000 | Gilad et al.[56] | SR | 43 | 43 | M | + | Ocular | 6 m | Normal | NR | + | LE | AChEI, PDN | Yes | 5 |
| 13 | 2001 | Musser et al.[48] | CR | 35 | 36 | M | + | Ocular | 1 y | Abnormal | + | NR | LE | Thymectomy, AChEI, MMF, PLEX | Yes | 5 |
| 14 |  |  |  | 10 | 28 | F | + | Ocular | 18 y | Abnormal | + | NR | UE | Thymectomy, AChEI, PDN, AZA, CsA | Yes |  |
| 15 | 2002 | Karacostas et al.[49] | LTE | 30 | 30 | F | + | Generalised | 0 | Abnormal | + | NR | UE | AChEI, PDN | Yes | 5 |
| 16 | 2003 | Scola et al.[57] | CR | 23 | 23 | F | + | Generalised | 7 y | Normal | + | + | LE | AChEI | Yes | 5 |
| 17 | 2006 | De Carvalho et al.[58] | LTE | 32 | 32 | F | + | Generalised | 0 | Normal | + | NR | UE | Thymectomy, AChEI, PDN, IVIG | Yes | 5 |
| 18 | 2007 | Samuraki et al.[50] | CR | 27 | 27 | M | + | Generalised | 0 | Abnormal | + | + | UE | Thymectomy, PLEX, IVIG, PDN | Yes | 5 |
| 19 | 2008 | Renard et al.[59] | CR | 17 | 24 | F | + | Generalised | 7 y | Normal | + | NR | UE | Thymectomy, PDN, IVIG | Yes | 5 |
| 20 | 2013 | Cirillo et al.[60] | CR | 62 | 62 | M | + | Generalised | 0 | Normal | + | NR | UE | AChEI, PDN | Yes | 5 |
| 21 | 2013 | Pardo et al.[61] | PA | 71 | 74 | M | + | Ocular | 3 y | NR | + | NR | UE | AChEI, PDN, IVIG | Yes | 5 |
| 22 | 2015 | Fearon et al.[51] | LTE | 39 | 39 | M | + | Generalised | 0 | Abnormal | + | NR | UE | AChEI, PDN, MMF, AZA, MTX, IVIG | Yes | 5 |
| 23 | 2016 | Andrade et al.[52] | PA | 29 | 47 | F | + | Generalised | 18 y | Abnormal | + | NR | UE | NR | NR | 5 |
| 24 | 2016 | Rodolico et al.[9] | RR | 22 | 33 | M | + | Ocular | 11y | NR | + | + | UE | AChEI, PDN | NR | 5 |
| 25 |  |  |  | 20 | 20 | F | + | Generalised | 4m | NR | + | NR | UE | Thymectomy, AChEI, PDN | NR |  |
| 26 |  |  |  | 40 | 40 | F | + | Generalised | 1m | NR | + | NR | UE | Thymectomy, AChEI, PDN | NR |  |
| 27 |  |  |  | 35 | 35 | M | + | Generalised | 6y | + | + | NR | UE | AChEI, PDN | NR |  |
| 28 | 2017 | Abraham et al.[10] | CS | 31 | 33 | M | + | Ocular | 2 y | NR | NR | + | UE | PDN, IVIG | Yes | 5 |
| 29 |  |  |  | 34 | 36 | M | - | Ocular | 2 y | NR | NR | + | UE | PDN, IVIG | Yes |  |
| 30 |  |  |  | 40 | 43 | M | + | Generalised | 3 y | NR | NR | + | UE | PDN, IVIG | Yes |  |
| 31 |  |  |  | 52 | 85 | F | + | Generalised | 33 y | Abnormal | + | + | UE | AChEI, MMF, PDN, IVIG | Yes |  |
| 32 |  |  |  | 55 | 58 | M | + | Ocular | 3 y | NR | + | + | UE | PDN, IVIG | Yes |  |
| 33 |  |  |  | 57 | 57 | M | - | Ocular | 6m | NR | + | + | UE | PDN, IVIG | Yes |  |
| 34 |  |  |  | 58 | 60 | M | + | Ocular | 2 y | NR | + | + | UE | PDN, IVIG | Yes |  |
| 35 | 2017 | Sousa et al.[47] | CR | 57 | 57 | M | - | Ocular | 6 w | Normal | + | NR | UE | AChEI, PDN, AZA | Yes | 5 |
| 36 | 2018 | Atmaca et al.[62] | CR | 61 | 61 | M | + | Generalised | 0 | NR | + | + | UE | AChEI,PDN | Yes | 5 |
| 37 | 2019 | Thilak et al.[11] | LTE | 47 | 47 | M | + | Generalised | 0 | NR | + | NR | UE | AChEI,PDN | Yes | 5 |
| 38 | 2021 | Mansukhani et al.[63] | CS | 53 | 53 | M | + | Generalised | 0 | NR | + | NR | UE | AChEI,PDN | Yes | 5 |
| 39 |  |  |  | 59 | 59 | M | + | Generalised | 0 | NR | + | NR | UE | AChEI,PDN | Yes |  |
| 40 |  |  |  | 34 | 34 | M | + | Generalised | 0 | NR | + | NR | UE | AChEI,PDN | Yes |  |
| 41 | 2021 | Shalabi et al.[64] | CR | 71 | 71 | F | + | Generalised | 0 | Normal | + | NR | UE | AChEI, PDN, PLEX | Yes | 5 |
| 42 | 2022 | Dragusin et al.[65] | CS | 64 | 64 | F | + | Generalised | 0 | Normal | + | NR | UE | AChEI,PDN | Yes | 5 |
| 43 | 2022 | Alanazy et al.[53] | CR | 29 | 35 | F | + | Generalised | 6 y | Abnormal | + | NR | UE | AChEI, PDN, IVIG, Rituximab | Yes | 5 |

Abbreviations: AChEI=Acetylcholinesterase Inhibitor; AcR-Ab=Acetylcholine Receptor Antibody; AZA=Azathioprine; BC=Brief Communications; CR=Case Report; CS=Case Series; CsA=Ciclosporin; CSN=Clinical/Scientific Note; EMG=Electromyography; F=Female; FU=Follow-up; IVIG=Intravenous Immunglobulin; LE=Lower Extremity; LTE=Letters to the Editor; m=months; M=Male; MG=Myasthenia Gravis; MMF=Mycophenolate Mofetil; MTX=Methotrexate; NR=Not Reported; OCEBM=Oxford Centre for Evidence-Based Medicine Level; PA=Poster Abstract; Pat. No.=Patient Number; PDN=Prednisone; PLEX=Plasmapheresis; Pred. W=Predominant Weakness; Pub.=Publication Type; Ref.=Reference; RNS=Repetitive Nerve Stimulation; RR=Research Report; SFEMG=Single Fiber EMG; SR=Brief Communications; Tx=Treatment; Tx Resp.=Treatment Response; UE=Upper Extremity; y=years; w=weeks;

**Table 2:**
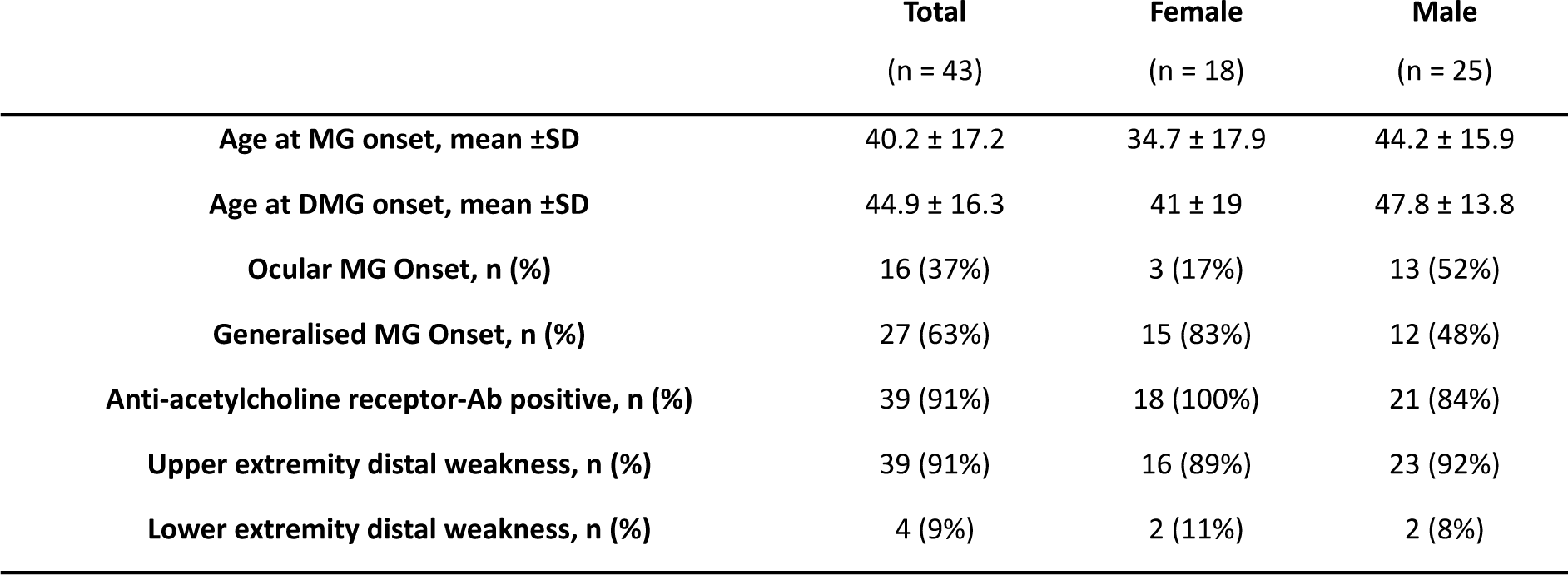
Summary of demographic and clinical characteristics of DMG patients included in this review.

|  | **Total**<br>(n = 43) | **Female**<br>(n = 18) | **Male**<br>(n = 25) |
| --- | --- | --- | --- |
| **Age at MG onset, mean ±SD** | 40.2 ± 17.2 | 34.7 ± 17.9 | 44.2 ± 15.9 |
| **Age at DMG onset, mean ±SD** | 44.9 ± 16.3 | 41 ± 19 | 47.8 ± 13.8 |
| **Ocular MG Onset, n (%)** | 16 (37%) | 3 (17%) | 13 (52%) |
| **Generalised MG Onset, n (%)** | 27 (63%) | 15 (83%) | 12 (48%) |
| **Anti-acetylcholine receptor-Ab positive, n (%)** | 39 (91%) | 18 (100%) | 21 (84%) |
| **Upper extremity distal weakness, n (%)** | 39 (91%) | 16 (89%) | 23 (92%) |
| **Lower extremity distal weakness, n (%)** | 4 (9%) | 2 (11%) | 2 (8%) |

Out of the 43 DMG patients included in the study, four tested negative for acetylcholine antibodies [10,46,47]. Further analysis showed that three of these individuals had positive single-fiber electromyography (EMG) results [10,46], all had typical clinical symptoms, exhibited a decremental response to repetitive nerve stimulation, and showed a positive response to treatment, which confirmed the diagnosis of myasthenia gravis [47]. However, we did exclude one case from one of the studies due to incomplete electrophysiological study, negative antibodies and a muscle biopsy indicating a myopathic pathology, that we felt required further evaluation [10].

During the review of the cases, we also observed that eight patients had abnormal electromyography (EMG) findings that resembled myopathy [10,48–52]. All those patients had a positive repetitive nerve stimulation (RNS) study. Out of the eight cases that showed abnormal EMG findings resembling myopathy, four cases had undergone muscle biopsies, which did not show myopathic abnormalities [10,50,52,53]. Among these four cases, two had single-fiber electromyography results that supported the diagnosis of myasthenia [10,50]. The four remaining cases that lacked muscle biopsy data were analysed thoroughly, and we considered several factors before including them in the study. These factors included clinical presentation, treatment response to cholinesterase agents, electrophysiological data, and blood parameters, such as a negative voltage-gated calcium channel antibody and a normal creatinine kinase level [48,49,51].

### 2.5. Quality Assessment of the articles

The two reviewers assessed the quality of the articles individually using the Oxford Centre for Evidence-Based Medicine Levels of Evidence [54]. Since all the articles were either case reports or case series, they were classified as OCEBM Level 5.

### 2.6. Data Analysis

We performed descriptive statistics of pooled individual patient data using R version 4.2.1, which we extracted from eligible case reports/series. The primary goal was to summarise and visualise this subgroup’s demographic and clinical features.

## 3. Results

Twenty-three publications consisting of 11 case reports, 5 letters to the editor, 2 poster abstracts and 5 case series fulfilled the inclusion and exclusion criteria. However, data from two case series could not be taken in totum [9,10]. In each of these two case series, one case was excluded due to incomplete data to rule out other possible causes of distal extremity weakness.

### 3.1. Demographic and Clinical Features

We considered a total of 43 MG patients with DMG. The mean age of onset for MG was 40.2 years (SD 17.2), while the mean age of onset for DMG was 44.9 years (SD 16.3). Among included DMG cases, men represented a slightly higher proportion (58%) than women (42%). Women were younger at MG and DMG onset, which was in concordance with the existing epidemiological data [2]. Most women with DMG (83%) presented with generalised MG at disease onset. However, in men the distribution was rather similar with 48% generalised MG and 52% ocular MG at disease onset. Across the whole group, a high percentage of patients (91%) tested positive for acetylcholine receptor antibodies, and this was true for all women in selected cases. Distal weakness was more common in the upper extremities (91%) than in the lower extremities for both sexes.

### 3.2. Reported time intervals from myasthenia onset to first distal weakness symptoms

One objective of our study is to analyse the duration between the onset of myasthenia and the initial manifestation of distal extremity weakness by examining individual case reports. To illustrate this, Fig. 2 presents the percentages of DMG patients and their time intervals from MG onset to first distal weakness, categorised by both sex and overall.

**Fig. 2:**
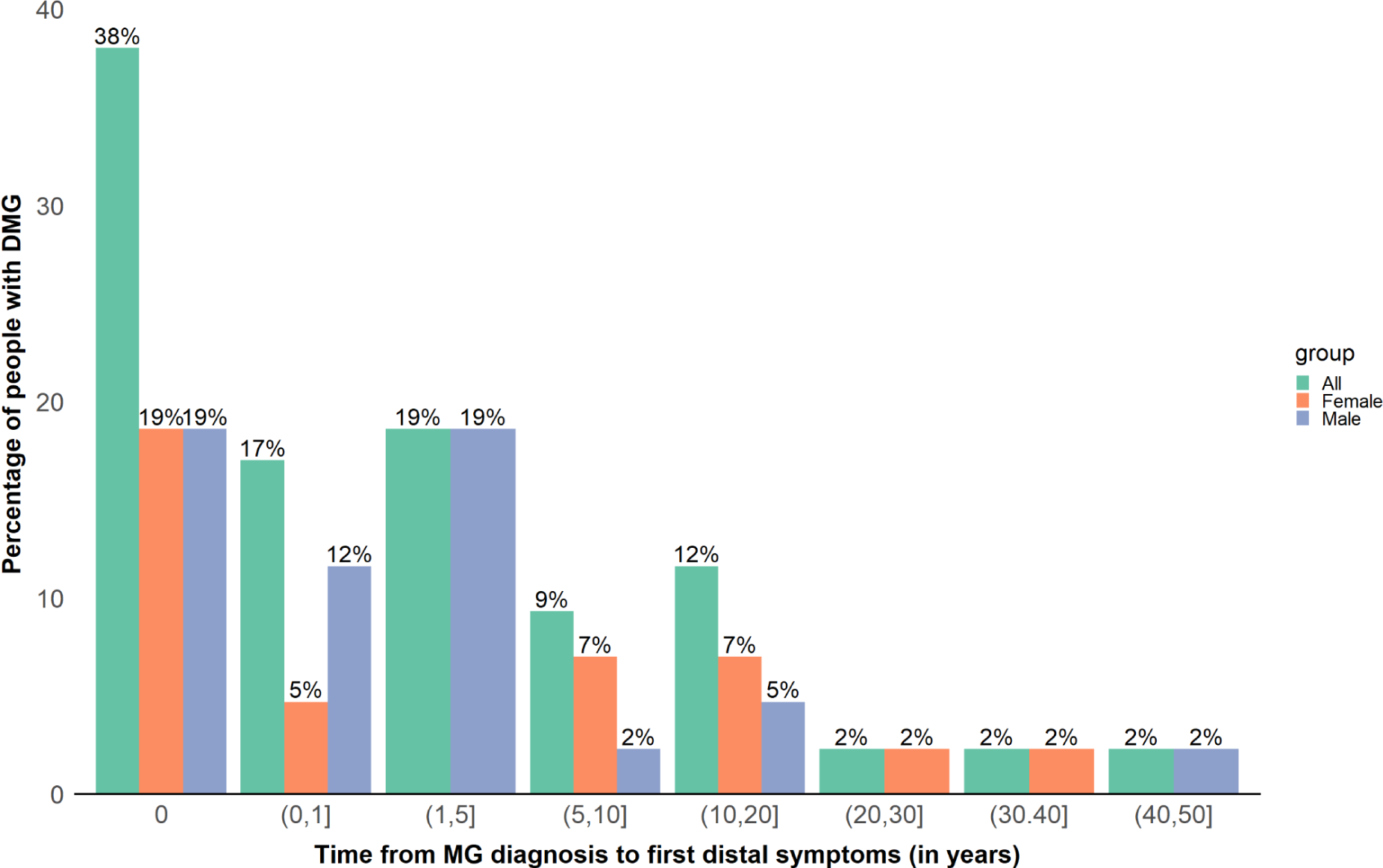
Temporal distribution of distal weakness onset in 43 DMG patients, categorised by the time since MG diagnosis: at onset (0), within 1 year (0,1]), 1-5 years (1,5]), 5-10 years (5,10]), 10-20 years (10,20]), 20-30 years (20,30]), 30-40 years (30,40]), and 40-50 years (40,50]).

Our results indicate that most patients (83%) experienced distal extremity weakness within ten years of their MG diagnosis. Furthermore, 38% of patients experienced distal extremity weakness at the onset of the disease, while 55% developed it within the first year of their MG. Notably, we observed one patient who had the longest interval between MG onset and the development of DMG, which was 45 years [6].

All reported data on Medical Research Council (MRC) scales regarding distal weakness were extracted from individual cases. In 3 patients, there was insufficient data regarding individual distal muscle groups (Patient No. 39, 40, 42) [63,65]. We extracted and considered the clinical features from the available case descriptions and visualised the distribution of weakness in different muscle groups for each patient, as well as the aggregate data in Figure 3.

**Fig. 3:**
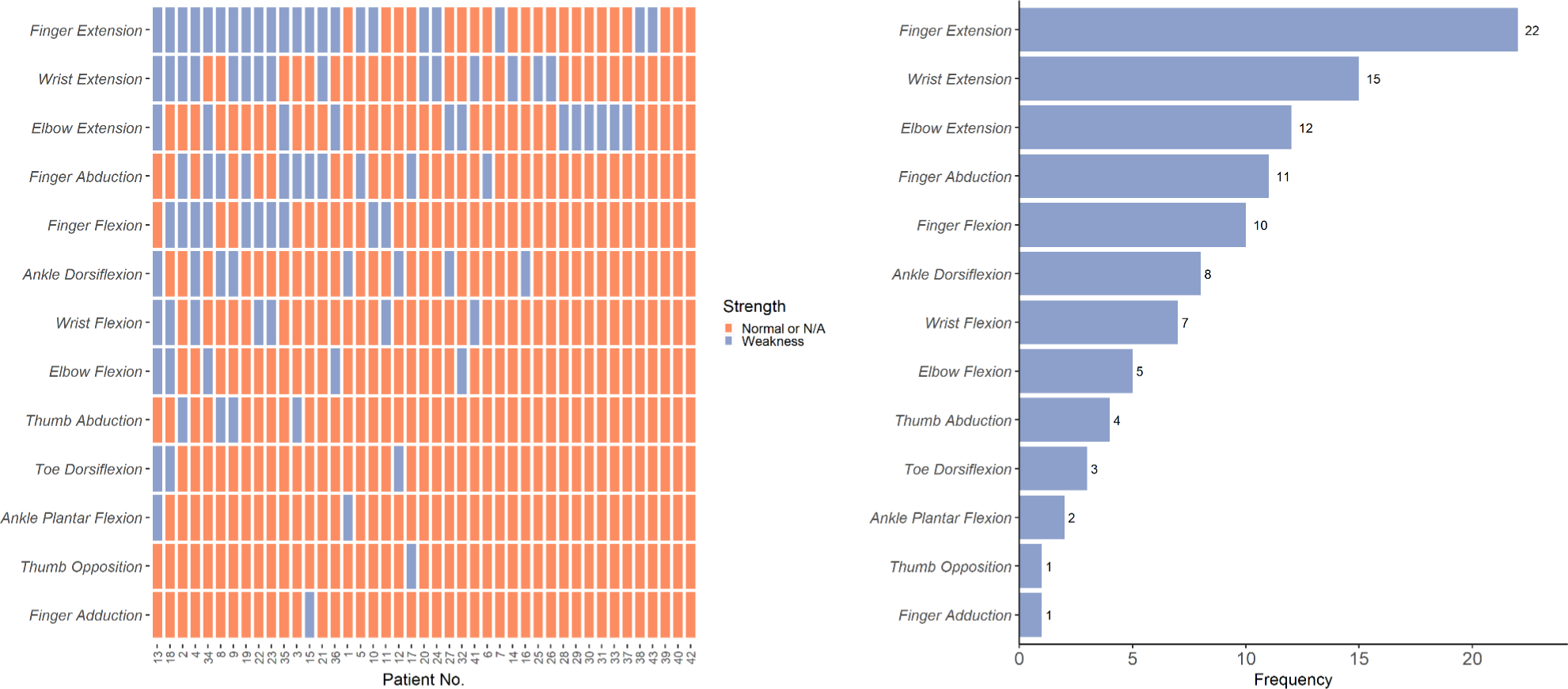
(Left) Distribution of distal weakness in DMG patients: The x-axis lists individual DMG patients numbers, while the y-axis indicates muscle function. For three patients (39, 40, 42), individual muscle data could not be extracted from the publications. Lilac denotes reported weakness, and orange signifies normal function or no report. **(Right) Frequency of reported distal muscle weakness:** Finger extension is the most common weakness, reported in 22 patients, followed by wrist extension weakness in 15 patients, elbow extension weakness in 12 patients.

Due to the lack of specific distal muscle weakness data for three patients, we report the cumulative muscle weaknesses based on 40 reported cases of distal weakness. Among these cases, finger extension was the most frequently reported type (22 cases), followed by wrist extension (15 cases), elbow extension (12 cases), finger abduction (11 cases), and finger flexion (10 cases). Ankle dorsiflexion weakness was the most commonly reported form of distal lower extremity weakness, appearing in eight cases.

## 4. Discussion

The number of clinical reports on distal weakness in MG appears to be slowly but steadily increasing, likely due to improved awareness. Since distal symptoms in MG are rare and unfamiliar to many health professionals, patients with MG who experience only distal extremity weakness at disease onset may face a diagnostic delay ranging from several weeks [47] to years [51]. According to a recent study, patients with generalised myasthenia gravis who experienced diagnosis delays of more than one year had more severe symptoms including higher rates of fatigue, anxiety, and depression, as well as a greater likelihood of advanced disease severity compared to those diagnosed within a year [66].

In our review of 43 DMG patients, the most common distal weaknesses involved finger and wrist extension in the upper limbs and ankle dorsiflexion in the lower limbs, with upper extremities being more frequently affected. This finding aligns with previously reported in MG cohorts with rather smaller numbers of DMG patients [5,6,49]. Notably, we found that approximately half exhibited distal weakness within the first year of MG diagnosis. Also a new finding showed that women with DMG might more often present with generalised MG at disease onset, a pattern which was not observed in men in our data.

Although patients initially presented with asymmetric weakness, they most eventually developed bilateral distal weakness, as also noted in previous studies [6]. Abnormal repetitive nerve stimulation (RNS) of the affected distal muscles was present in most patients, although data on RNS were insufficient in some cases [6,10,56]. Fatigability of distal muscles was documented in some DMG patients [11,56], but not in others [50,52,60], clearly.

DMG has an impact on daily functions and the quality of life of patients. In our selected cases, we observed that distal weakness often resulted in difficulties with fine motor skills such as doing up buttons [46,49,63], writing[46], using cutlery[46], counting money[63], plaiting hair[63], holding objects [49,63], sewing[58], using clothes pegs [58] and walking [55,56].

Although treatment response was generally positive across most cases, some reports lacked clinical information to draw clear conclusions [6,9,52]. In cases where it was reported, patients generally responded well to treatment and achieved good remission. However, in one case with initially severe finger extension weakness (MRC 0/5), there was incomplete remission after 2 years of treatment (MRC 3/5) [53]. This suggests that prompt and accurate diagnosis, along with potentially aggressive treatment, may be necessary in some severe cases of distal weakness associated with MG.

In standardised scales such as the manual muscle test (MMT) for MG [67], MRC sum-score[68] and Quantitative Myasthenia Gravis (QMG) [69], distal muscles, especially finger extension function, are not completely represented. The insufficient focus on distal muscles in these scales could pose challenges in identifying distal symptoms of MG. Neglecting to address these symptoms in assessments may sequentially have negative consequences on patients’ quality of life. Therefore, there is a need to improve the current standardised scales to better apprehend distal muscle involvement in MG. Patients with MG could benefit from more frequent measurements using electronic devices, which can collect objective digital biomarkers and document daily fluctuations in distal extremity function in the near future.

### 4.1. Differential diagnoses for distal weakness in MG

When patients present with symptoms of distal weakness, it is important to conduct a thorough evaluation before concluding that it is caused by myasthenia gravis. Other potential causes to consider include disorders of the upper motor neurons such as lesions in the brain and spine, disorders involving anterior horn cells like motor neuron disease or spinal atrophy, peripheral nerve disorders like mono- and polyneuropathies, and other neuromuscular junction disorders such as congenital myasthenia and Lambert-Eaton Myasthenic Syndrome (LEMS). Additionally, mitochondrial myopathies and hereditary distal myopathies should be included in the differential diagnosis [70,71].

When evaluating a patient with distal weakness and suspicion of myasthenia gravis (MG), we suggest considering two MG-associated conditions with possible distal extremity weakness. The first one is the myasthenia-inflammatory myopathy (MG-IM) overlap syndrome and the second one is myasthenic neuromyopathy.

#### 4.1.1. Myasthenia-Inflammatory Myopathy (MG-IM) Overlap Syndrome

MG-IM is characterised by non-fluctuating muscle weakness that worsens progressively over a period of weeks or months and is often accompanied by myalgias [72]. It is a rare condition that affects less than 3% of myasthenic patients. It can present as polymyositis (63%), dermatomyositis (25%), or granulomatosis (12%) [72]. Patients with myositis may exhibit systemic symptoms such as interstitial lung disease, arthritis/arthralgia, Raynaud’s phenomenon, and skin lesions that are not typically seen in MG. In addition, myositis-specific (MSA) and myositis-associated autoantibodies (MAA) and an elevated serum Creatinine Kinase (CK) are features seen in IM which are not found in MG patients [72]. Skeletal muscle magnetic resonance imaging (MRI) in patients with IM may show signs of edema and inflammation in short tau inversion recovery (STIR), sometimes associated with fibro-fatty replacement in T1 sequences [73].

The neurophysiological hallmarks in MG patients include a decremental response of compound motor action potential (CMAP) during repetitive nerve stimulation (RNS) and increased jitter and blocks during single-fiber electromyography (SFEMG). However, electromyography may show an additional myogenic pattern such as spontaneous activity and SASD (small amplitude short duration) motor unit action potentials in MG-IM patients [74]. Despite these differences, muscle biopsy remains the gold standard for diagnosis and should be considered in all cases with high suspicion of MG-IM[75].

#### 4.1.2. Myasthenic Neuromyopathy

Myasthenic neuromyopathy refers to the presence of myopathic changes on electromyography (EMG) with or without muscle atrophy in patients with myasthenia gravis (MG) [76]. In 1958, Ossermann et al. described a subset of MG patients as Group V, who initially presented with generalised myasthenia and began to show muscle atrophy as early as six months after onset [77]. Kinoshita et al. conducted a study on a similar group of patients with myasthenia gravis (MG) with significant muscle atrophy, myopathic EMG changes, and elevated creatine kinase (CK) levels. These patients responded well to anticholinesterase therapy and their muscle biopsies showed no signs of inflammation that would indicate myositis [76].

Similarly, Samuraki et al. reported a case with probable myasthenic neuromyopathy. This patient had normal CK levels, but electrophysiological studies revealed myopathic changes. In addition, the muscle biopsy did not show signs of myositis but did reveal decreased reactivity to acetylcholine receptors (*AChR*) and deposits of complement C3 commonly found in MG [50].

According to previous pathology studies, such as the one conducted by Fenichel et al. [75], myasthenia gravis might cause both neurogenic and myopathic changes over time. However, these changes are typically mild, and fasciculations are rather uncommon [75]. The myopathic changes are likely secondary to denervation in MG, but further research is needed to fully understand this phenomenon and its underlying mechanisms.

### 4.2. Recommendations regarding design of prospective studies on DMG

After reviewing previous literature on DMG and analysing the excluded data from our study, we have compiled a list of recommendations for researchers planning prospective studies on DMG depicted in Table 3.

**Table 3:**
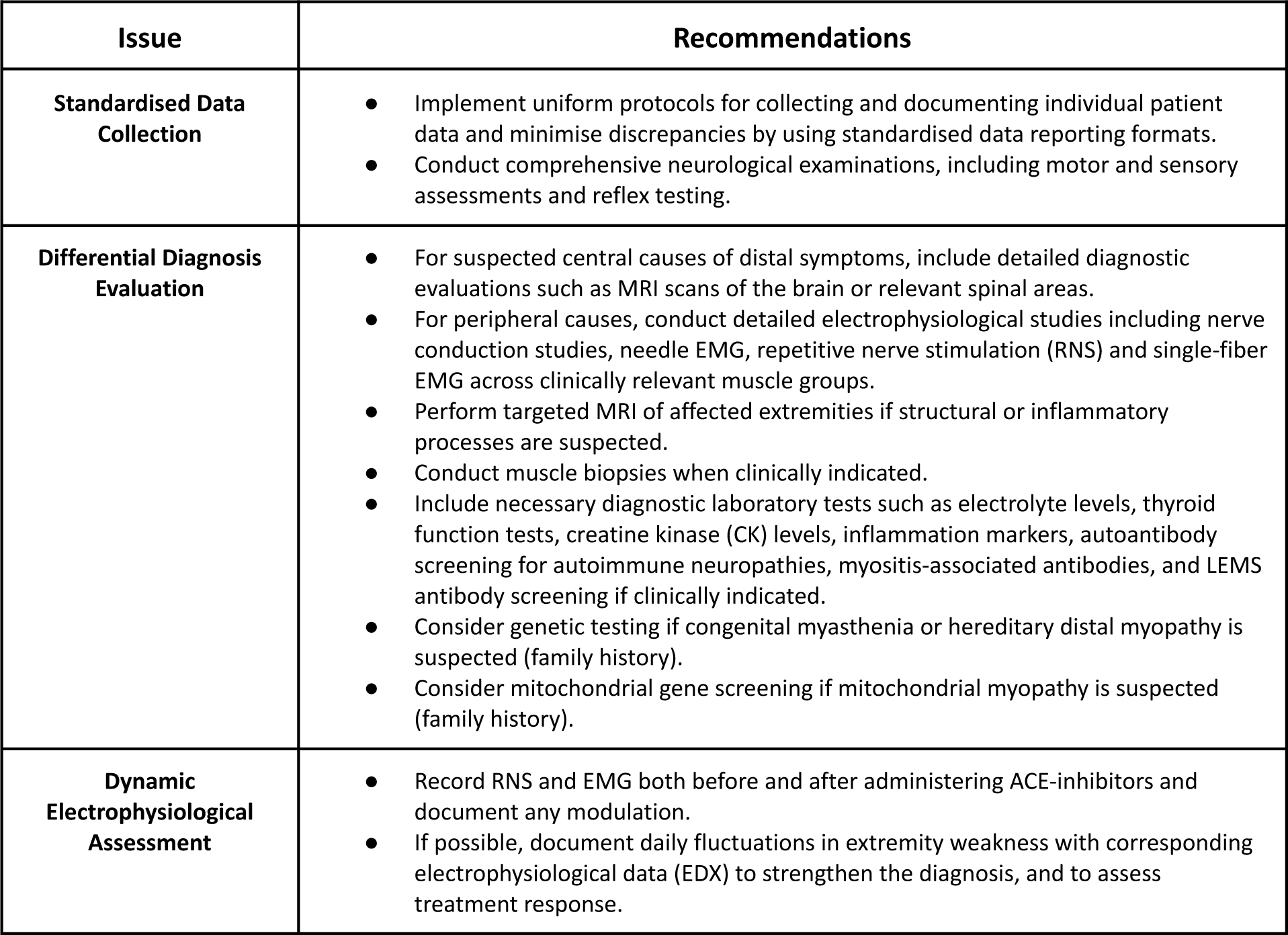
Recommendations for prospective studies on DMG.

| Issue | Recommendations |
| --- | --- |
| **Standardised Data Collection** | • Implement uniform protocols for collecting and documenting individual patient data and minimise discrepancies by using standardised data reporting formats.<br>• Conduct comprehensive neurological examinations, including motor and sensory assessments and reflex testing. |
| **Differential Diagnosis Evaluation** | • For suspected central causes of distal symptoms, include detailed diagnostic evaluations such as MRI scans of the brain or relevant spinal areas.<br>• For peripheral causes, conduct detailed electrophysiological studies including nerve conduction studies, needle EMG, repetitive nerve stimulation (RNS) and single-fiber EMG across clinically relevant muscle groups.<br>• Perform targeted MRI of affected extremities if structural or inflammatory processes are suspected.<br>• Conduct muscle biopsies when clinically indicated.<br>• Include necessary diagnostic laboratory tests such as electrolyte levels, thyroid function tests, creatine kinase (CK) levels, inflammation markers, autoantibody screening for autoimmune neuropathies, myositis-associated antibodies, and LEMS antibody screening if clinically indicated.<br>• Consider genetic testing if congenital myasthenia or hereditary distal myopathy is suspected (family history).<br>• Consider mitochondrial gene screening if mitochondrial myopathy is suspected (family history). |
| **Dynamic Electrophysiological Assessment** | • Record RNS and EMG both before and after administering ACE-inhibitors and document any modulation.<br>• If possible, document daily fluctuations in extremity weakness with corresponding electrophysiological data (EDX) to strengthen the diagnosis, and to assess treatment response. |

Distal extremity weakness can be attributed to various diseases and aetiologies, which might make systematic tracking in DMG patients challenging. To address this, we recommend prioritising the clinical examination and its detailed description by including a comprehensive motor and sensory examination with reflex testing. Depending on additional clinical markers and impressions, further investigation may include more extensive assessments.

### 4.3. Limitations of the review and future perspectives

This scoping review has two significant limitations. Firstly, the number of studies included is relatively small, and the literature primarily consists of case reports and case series. While case reports can be valuable in identifying rare presentations, they are anecdotal and cannot guarantee completeness. Secondly, a standardised reporting of MRC scales for examining distal muscle groups lacked across the analysed studies, which probably have influenced our results.

Despite these limitations, this study provides new insights into the DMG population. However, studies with larger sample sizes and standardised protocols for data collection and analysis are needed to enhance the quality of future research.

## 5. Conclusion

Current evidence suggests that distal extremity weakness occurs in a subgroup of MG patients and may be more prevalent in MG than previously believed. To enhance our understanding of this aspect of MG, prospective studies are needed, particularly with regards to DMG’s aetiology and its optimal treatment strategies. Future research should also explore the efficacy of current treatments for DMG and investigate its impact on quality of life.

## Data Availability

All data produced in the present work are contained in the manuscript.

## Abbreviations

AChR: Acetylcholine receptor
DMG: Distal myasthenia gravis
EMG: Electromyography
MG: Myasthenia gravis
OCEBM: Oxford Centre for Evidence-Based Medicine
RNS: Repetitive nerve stimulation
MSA: Myositis-specific autoantibodies
MAA: Myositis-associated autoantibodies
CK: Creatine kinase
MG-IM: Myasthenia gravis with inflammatory myopathy
LEMS: Lambert-Eaton myasthenic syndrome
QMGS: Quantitative myasthenia gravis score

## Acknowledgments

We extend our heartfelt gratitude to Carolyn Smith of the Oxford Bodleian Library for her continous support throughout this scoping review. Additionally, we sincerely thank Dr. Ross Patrick Byrne from Trinity College Dublin for his valuable insights and guidance on the text’s structure. Finally, E.K. expresses his appreciation to the MND Association for providing his current PhD funding (MND Association 979-799).

## References

1. Vincent A, Huda S, Cao M, Cetin H, Koneczny I, Rodriguez-Cruz P, et al. Serological and experimental studies in different forms of myasthenia gravis. Ann N Y Acad Sci. 2018 Feb;1413[1]:143–53. doi:10.1111/nyas.13592.

2. Carr AS, Cardwell CR, McCarron PO, McConville J. A systematic review of population based epidemiological studies in Myasthenia Gravis. BMC Neurol. 2010 Jun 18;10:46. doi:10.1186/1471-2377-10-46.

3. Carey IM, Banchoff E, Nirmalananthan N, Harris T, DeWilde S, Chaudhry UAR, et al. Prevalence and incidence of neuromuscular conditions in the UK between 2000 and 2019: A retrospective study using primary care data. PLoS ONE. 2021 Dec 31;16[12]:e0261983. doi:10.1371/journal.pone.0261983.

4. Verschuuren JJGM, Palace J, Erik Gilhus N. Clinical aspects of myasthenia explained. Autoimmunity. 2010 Aug;43[5–6]:344–52. doi:10.3109/08916931003602130.

5. Harvey AM. Some Preliminary Observations on the Clinical Course of Myasthenia Gravis before and after Thymectomy. Bull N Y Acad Med. 1948 Aug;24[8]:505–22.

6. Nations SP, Wolfe GI, Amato AA, Jackson CE, Bryan WW, Barohn RJ. Distal myasthenia gravis. Neurology. 1999 Feb 1;52[3]:632–632. doi:10.1212/WNL.52.3.632.

7. Werner P, Kiechl S, Löscher W, Poewe W, Willeit J. Distal myasthenia gravis - frequency and clinical course in a large prospective series. Acta Neurol Scand. 2003 Sep;108[3]:209–11. doi:10.1034/j.1600-0404.2003.00136.x.

8. Fitzgerald MG, Shafritz AB. Distal myasthenia gravis. J Hand Surg. 2014;39[7]:1419–20. doi:10.1016/j.jhsa.2013.12.036.

9. Rodolico C, Parisi D, Portaro S, Biasini F, Sinicropi S, Ciranni A, et al. Myasthenia Gravis: Unusual Presentations and Diagnostic Pitfalls. J Neuromuscul Dis. 2016;3[3]:413–8. doi:10.3233/JND-160148.

10. Abraham A, Kassardjian CD, Katzberg HD, Bril V, Breiner A. Selective or predominant triceps muscle weakness in African–American patients with myasthenia gravis. Neuromuscul Disord. 2017 Jul 1;27[7]:646–9. doi:10.1016/j.nmd.2017.04.009.

11. Thilak MR, Prabhu AN, Sai Rao KS, Vasantbhai MJ. Isolated bilateral triceps muscle weakness as a presenting complaint in myasthenia gravis: A review. Neurol India. 2019 Mar 1;67[2]:566. doi:10.4103/0028-3886.258004.

12. Narayanaswami P, Sanders DB, Bibeau K, Krueger A, Venitz J, Guptill JT. Identifying a patient-centered outcome measure for a comparative effectiveness treatment trial in myasthenia gravis. Muscle Nerve. 2022 Jan;65[1]:75–81. doi:10.1002/mus.27443.

13. Barnett C, Herbelin L, Dimachkie MM, Barohn RJ. Measuring Clinical Treatment Response in Myasthenia Gravis. Neurol Clin. 2018 May 1;36[2]:339. doi:10.1016/J.NCL.2018.01.006.

14. Peters MDJ, Godfrey C, McInerney P, Munn Z, Tricco AC, Khalil H. Chapter 11: Scoping Reviews (2020 version). In: Aromataris E, Munn Z, editors. JBI Manual for Evidence Synthesis. JBI; 2020.

15. Sussman J, Farrugia ME, Maddison P, Hill M, Leite MI, Hilton-Jones D. Myasthenia gravis: Association of British Neurologists’ management guidelines. Pract Neurol. 2015 Jun 1;15[3]:199–206. doi:10.1136/practneurol-2015-001126.

16. Ouzzani M, Hammady H, Fedorowicz Z, Elmagarmid A. Rayyan—a web and mobile app for systematic reviews. Syst Rev [Internet]. 2016 [cited 2022 Aug 9];5[1]. Available from: https://link.springer.com/epdf/10.1186/s13643-016-0384-4doi:10.1186/s13643-016-0384-4.

17. Vellodi C, Tallis RC. An Unusual Case of Myasthenia gravis in an Elderly Patient with Severe Muscular Atrophy. Gerontology. 2009 Apr 7;34[4]:209–11. doi:10.1159/000212955.

18. Mazia CG, Schottlender J, Herrera M, Menga G, Rey L, Rivero A. Bilateral foot drop as initial isolated manifestation of distal myasthenia [Abstract]. Neuromuscul Disord. 1999 Oct 1;9[6]:474–534. doi:10.1016/S0960-8966(99)00090-5.

19. Öztürk A, Deymeer F, Serdaroglu P, Gulsen-Parman Y, Özdemir C. Distribution of muscle weakness in myasthenia gravis [Abstract]. Neuromuscul Disord. 1999 Oct 1;9[6]:474–534. doi:10.1016/S0960-8966(99)00090-5.

20. Deymeer F, Serdaroglu P, Parman Y, Kiliç A, Özdemir C. Onset with limb weakness in myasthenia gravis: frequent presentation with leg weakness in the second decade. Neuromuscul Disord. 2003 Sep;13[7–8]:662–662.

21. Iwasaki Y, Igarashi O, Kawabe K, Kiyozuka T, Kawase Y, Aoyagi J, et al. MG with distal muscle involvement. Acta Neurol Scand. 2004 Oct;110[4]:270–270. doi:10.1111/j.1600-0404.2004.00312.x.

22. Nicolle MW. Wrist and Finger Drop in Myasthenia Gravis. J Clin Neuromuscul Dis. 2006 Dec;8[2]:65–9. doi:10.1097/01.cnd.0000245218.70927.97.

23. Youn JY, Kim MK, Bae JS. Finger extensor weakness in myasthenia gravis patients. Neuromuscul Disord. 2006 Jul;16:S117–S117.

24. Wee AS, Subramony SH. PO26-TH-23 Wrist drop mimicking radial nerve palsy in myasthenia gravis. J Neurol Sci. 2009 Oct;285:S305. doi:10.1016/S0022-510X(09)71164-5.

25. Nikolic DM, Nikolic AV, Lavrnic DV, Medjo BP, Ivanovski PI. Childhood-Onset Myasthenia Gravis With Thymoma. Pediatr Neurol. 2012 May;46[5]:329–31. doi:10.1016/j.pediatrneurol.2012.02.025.

26. Misra I, Temesgen FD, Soleiman N, Kalyanam J, Kurukumbi M. Myasthenia gravis presenting like guillain-barré syndrome. Case Rep Neurol. 2012;4[3]:137–43. doi:10.1159/000342448.

27. Zouvelou V, Zisimopoulou P, Psimenou E, Matsigkou E, Stamboulis E, Tzartos SJ. AChR-myasthenia gravis switching to double-seropositive several years after the onset. J Neuroimmunol. 2014 Feb 15;267[1]:111–2. doi:10.1016/j.jneuroim.2013.12.012.

28. Merino-Ramírez MA. ID 209 – Uncommon variants of Myasthenia Gravis clinical onset in a large retrospective series. Clin Neurophysiol. 2016 Mar;127[3]:e89–90. doi:10.1016/j.clinph.2015.11.300.

29. Florendo-Cumbermack AG, Nicolle MW. B.04 Distal and asymmetric myasthenia gravis: a case series of 54 patients. Can J Neurol Sci. 2016 Jun;43[S2]:S10–S10. doi:10.1017/cjn.2016.63.

30. Nakanishi T, Hamada O, Nakai M. A Guillain-Barre Syndrome Mimicry, Myasthenia Gravis. Diagnosis. 2017 Nov 27;4[4]:eA43–124. doi:10.1515/dx-2017-0034.

31. Masselli R. Hand Myasthenia in a patient with positive acetylcholine receptor antibodies. Muscle Nerve. 2019;60[S1]:S1–140. doi:10.1002/mus.26647.

32. Missel MF, Young J. Poster A weak suspicion revealed the reason for a patients weakness: An atypical presentation of myasthenia gravis. J Gen Intern Med. 2020 Jul;35[S1]:1–779. doi:10.1007/s11606-020-05890-3.

33. Ferro M. Distal Presenting Myasthenia Gravis Eletrophysiologic Tests Role. Eur J Neurol. 2021;28[S1]:442–921. doi:10.1111/ene.14975.

34. Sotero FD, Castro I, Santos MO. Focal and asymmetric triceps brachii weakness: a rare but possible myasthenia gravis presentation. Neurol Sci. 2022 Apr;43[4]:2871–3. doi:10.1007/s10072-022-05886-3.

35. Özenç B, Işık K, Odabaşı Z. Isolated Bilateral Triceps Weakness in Myasthenia Gravis. Acta Neurol Taiwanica. 2024 Jun 30;33(2):73–5.

36. Alhabib A, Hashim H, Hussain Y, Barrientos M, Gonsoulin S, Guilbeau L. Unusual Etiology of Bilateral Wrist Drop. J Clin Neuromuscul Dis. 2023;24[(Alhabib A.; Hashim H.; Hussain Y.; Barrientos M.; Gonsoulin S.; Guilbeau L.) Austin, TX, United States]:S4–5.

37. Chancellor K. Rare Presentation of Myasthenia Gravis: a case study. South Med J. 2023;116[7]:623–4.

38. Lalive PH, Allali G, Truffert A. Myasthenia gravis associated with HTLV-I infection and atypical brain lesions. Muscle Nerve. 2007 Apr;35[4]:525–8. doi:10.1002/mus.20694.

39. Rubin DI, Litchy WJ. Severe, focal tibialis anterior and triceps brachii weakness in myasthenia gravis: A case report. J Clin Neuromuscul Dis. 2011 Jun;12[4]:219–22. doi:10.1097/CND.0B013E3181DC7C5B.

40. Deters D, Fowler SL, Orozco R, Smith PR, Spurlock S, Blackmon D, et al. Myasthenia gravis presentation after a cervical laminectomy with fusion. Dimens Crit Care Nurs. 2016 Jul 1;35[4]:190–4. doi:10.1097/DCC.0000000000000189.

41. Domingo CA, Landau ME, Campbell WW. Selective Triceps Muscle Weakness in Myasthenia Gravis is Under-Recognized. J Clin Neuromuscul Dis. 2016 Dec;18[2]:103–4. doi:10.1097/CND.0000000000000121.

42. Ng JL, Hooi WF, Malhotra A, Talman P. Myasthenia gravis with an unusual pattern of weakness and positive igg anti gm1 antibody. J Neurol Neurosurg Psychiatry. 2017 May 1;88[5]:e1–e1. doi:10.1136/jnnp-2017-316074.27.

43. Chaudhry V, Corse A, Weiner L, Drachman D. Foot drop in myasthenia gravis. NEUROLOGY. 1998 Apr;50[4]:A82–3.

44. Nicolle M, Watson B. Myasthenia gravis presenting with distal asymmetric weakness. MUSCLE NERVE. 2005 Sep;32[3]:417–417.

45. Jian F, Wang HB, Chen N, Yang S, Liu Y, Zhao YZ, et al. Observation of clinical and electrophysiological features in patients with distal myasthenia gravis. Zhonghua Yi Xue Za Zhi. 2017 Oct 10;97:2894–7. doi:10.3760/cma.j.issn.0376-2491.2017.37.004.

46. Janssen JC, Larner AJ, Harris J, Shean GL, Rossor MN. Myasthenic hand. Neurology. 1998 Sep 1;51[3]:913–4. doi:10.1212/WNL.51.3.913.

47. Sousa DC, Viana P, Leal I, Parreira S, Ferreira J, Campos F. Ophthalmoparesis and unilateral finger flexor muscle weakness in seronegative myasthenia gravis. Can J Ophthalmol. 2017 Dec 1;52[6]:e213–6. doi:10.1016/J.JCJO.2017.04.012.

48. Musser WS, Barbano RL, Thornton CA, Moxley RT, Herrmann DN, Logigian EL. Distal myasthenia gravis with a decrement, an increment, and denervation. J Clin Neuromuscul Dis. 2001 Sep 1;3[1]:16–9. doi:10.1097/00131402-200109000-00004.

49. Karacostas D, Mavromatis I, Georgakoudas G, Artemis N, Milonas I. Isolated distal hand weakness as the only presenting symptom of myasthenia gravis. Eur J Neurol. 2002 Jul;9[4]:429–30. doi:10.1046/j.1468-1331.2002.00417.x.

50. Samuraki M, Furui E, Komai K, Takamori M, Yamada M. Myasthenia gravis presenting with unusual neurogenic muscle atrophy. Muscle Nerve. 2007;36[3]:394–9. doi:10.1002/mus.20757.

51. Fearon C, Mullins G, Reid V, Smyth S. Distal myasthenia gravis presenting as isolated distal myopathy. Muscle Nerve. 2015;52[2]:308–9. doi:10.1002/mus.24650.

52. Andrade R. Distal Myasthenia Gravis: An unusual presentation with slowly progressive distal weakness and atrophy. J Neuromuscul Dis. 2016 Jun 21;3[s1]:S1–231. doi:10.3233/JND-160001.

53. Alanazy MH, Alkhalidi H. Finger Flexor Weakness in Myasthenia Gravis. 2022;0.

54. OCEBM Levels of Evidence Working Group. The Oxford 2011 Levels of Evidence. Oxford Centre for Evidence-Based Medicine. [Internet]. [cited 2022 Aug 5]. Available from: https://www.cebm.ox.ac.uk/resources/levels-of-evidence/ocebm-levels-of-evidence

55. Uncini A, Di Guglielmo G, Di Muzio A, Repaci M, Lugaresi A, Forno GM, et al. Distal myasthenia gravis and sensory neuronopathy with anti-50 kDa antibody mimicking sensory-motor neuropathy. J Neurol Neurosurg Psychiatry. 1997 Sep 1;63[3]:414–414. doi:10.1136/JNNP.63.3.414.

56. Gilad R, Sadeh M. Bilateral Foot Drop as a Manifestation of Myasthenia Gravis: J Clin Neuromuscul Dis. 2000 Sep;2[1]:23. doi:10.1097/00131402-200009000-00006.

57. Scola RH, Iwamoto FM, Mainardi MAM, Della-Coletta MV, Carvalho G, Zavala JA, et al. Miastenia grave distal: relato de caso. Arq Neuropsiquiatr. 2003 Mar;61[1]:119–20. doi:10.1590/S0004-282X2003000100024.

58. de Carvalho M, Geraldes R. Longstanding right-hand weakness in a patient with myasthenia gravis. Muscle Nerve. 2006 Nov;34[5]:670–1. doi:10.1002/mus.20617.

59. Renard D, Castelnovo G, Labauge P. Distal myasthenia gravis. Acta Neurol Belg. 2008;108[3]:107–8.

60. Cirillo G, Todisco V, Tessitore A, Tedeschi G. Clinical reasoning: A 62-year-old man with right wrist drop. Neurology. 2013 Sep 10;81[11]. doi:10.1212/WNL.0B013E3182A43B90.

61. Pardo J, García-Sobrino T, Vidal MP, Pardo-Parrado M, Fernández-Pajarín G, Santamaría-Cadavid M. Distal myasthenia simulating radial nerve palsy: A case report. J Neurol Sci. 2013 Oct;333:e449. doi:10.1016/j.jns.2013.07.1605.

62. Atmaca MM, Durmus Tekce H, Kocasoy Orhan E, Baslo MB, Deymeer F. Myasthenia gravis with clinical facilitation: a case series. Neurol Sci Neurophysiol. 2018 Dec 5;35[4]:193–7. doi:10.5152/NSN.2018.11208.

63. Anil Mansukhani K, Sharma A, Balakrishnan L, Chavan P. Distal myasthenia gravis – A missed clinical diagnosis. Clin Neurophysiol. 2021;132[8]:e91–2. doi:10.1016/j.clinph.2021.02.206.

64. Shalabi F, Karussis D. A Unique Case of Unilateral Distal Arm Weakness as a Presentation of Myasthenia Gravis. Austin J Clin Neurol. 2021 Oct 4;8[3]. doi:10.26420/austinjclinneurol.2021.1153.

65. Dragusin A, Grecu N, Ribigan AC, Badea RS, Terecoasa EO, Ene A, et al. Low Fluctuation of Symptoms May Delay Diagnosis of Myasthenia Gravis: A Case Series. Neurol Ther. 2022 Mar 1;11[1]:481–7. doi:10.1007/s40120-021-00312-w.

66. Cortés-Vicente E, Borsi AJ, Gary C, Noel WGJ, Lee JMS, Karmous W, et al. The impact of diagnosis delay on European patients with generalised myasthenia gravis. Ann Clin Transl Neurol [Internet]. [cited 2024 Aug 11];n/a[n/a]. Available from: https://onlinelibrary.wiley.com/doi/abs/10.1002/acn3.52122doi:10.1002/acn3.52122.

67. Sanders DB, Tucker-Lipscomb B, Massey JM. A simple manual muscle test for myasthenia gravis: Validation and comparison with the QMG score. Ann N Y Acad Sci. 2003;998:440–4. doi:10.1196/ANNALS.1254.057.

68. Kleyweg RP, van der Meché FG, Schmitz PI. Interobserver agreement in the assessment of muscle strength and functional abilities in Guillain-Barré syndrome. Muscle Nerve. 1991 Nov;14[11]:1103–9. doi:10.1002/mus.880141111.

69. Sharshar T, Chevret S, Mazighi M, Chillet P, Huberfeld G, Berreotta C, et al. Validity and reliability of two muscle strength scores commonly used as endpoints in assessing treatment of myasthenia gravis. J Neurol. 2000 Apr;247[4]:286–90. doi:10.1007/s004150050585.

70. Savarese M, Sarparanta J, Vihola A, Jonson PH, Johari M, Rusanen S, et al. Panorama of the distal myopathies. Acta Myol. 2020 Dec 1;39[4]:245–65. doi:10.36185/2532-1900-028.

71. Beecher G, Gavrilova RH, Mandrekar J, Naddaf E. Mitochondrial myopathies diagnosed in adulthood: clinico-genetic spectrum and long-term outcomes. Brain Commun. 2024 Apr 1;6[2]:fcae041. doi:10.1093/braincomms/fcae041.

72. Garibaldi M, Fionda L, Vanoli F, Leonardi L, Loreti S, Bucci E, et al. Muscle involvement in myasthenia gravis: Expanding the clinical spectrum of Myasthenia-Myositis association from a large cohort of patients. Autoimmun Rev. 2020 Apr 1;19[4]:102498. doi:10.1016/j.autrev.2020.102498.

73. Grande FD, Carrino JA, Grande MD, Mammen AL, Stine LC. Magnetic resonance imaging of inflammatory myopathies. Top Magn Reson Imaging. 2011 Apr;22[2]:39–43. doi:10.1097/RMR.0b013e31825b2c35.

74. Paganoni S, Amato A. Electrodiagnostic Evaluation of Myopathies. Phys Med Rehabil Clin N Am. 2013;24[1]:193–207. doi:10.1016/j.pmr.2012.08.017.

75. Fenichel GM. Muscle Lesions in Myasthenia Gravis. Ann N Y Acad Sci. 1966;135[1]:60–7. doi:10.1111/j.1749-6632.1966.tb45463.x.

76. Kinoshita M, Nakazato H, Wakata N, Satoyoshi E. Myasthenic Neuromyopathy. Eur Neurol. 1982;21[1]:52–8. doi:10.1159/000115454.

77. Osserman Ke, Kornfeld P, Cohen E, Genkins G, Mendelow H, Goldberg H, et al. Studies in Myasthenia Gravis: Review of Two Hundred Eighty-Two Cases at The Mount Sinai Hospital, New York City. AMA Arch Intern Med. 1958 Jul 1;102[1]:72–81. doi:10.1001/archinte.1958.00260190074008.

