## Supplemental_File for "Clinical and Demographic Features of Distal Extremity Weakness in Myasthenia Gravis - A Scoping Review"

### S1 Appendix - Search strategy

**Databases searched:** MEDLINE, EMBASE, Scopus, Web of Science

**Grey Literature:** ResearchGate

**Date last search was run:** 08.08.2024

**Years covered by search:** All years from inception.

**Document restriction:** No restriction

#### Summary of the search strategy:

##### MEDLINE (Ovid) (557)

1 myasthenia.mp. [mp=title, abstract, heading word, drug trade name, original title, device manufacturer, drug manufacturer, device trade name, keyword heading word, floating subheading word, candidate term word]

2 (distal or arm or wrist or hand or finger or thumb or leg or ankle or foot or triceps or biceps or tibialis).mp. [mp=title, abstract, heading word, drug trade name, original title, device manufacturer, drug manufacturer, device trade name, keyword heading word, floating subheading word, candidate term word]

3 1 and 2

##### EMBASE (Embase.com) (1110)

1 myasthenia:kw,ab,ta,ti,tn,mn,jt,pt

2 distal:kw,ab,ta,ti,tn,mn,jt,pt OR arm:kw,ab,ta,ti,tn,mn,jt,pt OR wrist:kw,ab,ta,ti,tn,mn,jt,pt OR hand:kw,ab,ta,ti,tn,mn,jt,pt OR finger:kw,ab,ta,ti,tn,mn,jt,pt OR thumb:kw,ab,ta,ti,tn,mn,jt,pt OR leg:kw,ab,ta,ti,tn,mn,jt,pt OR ankle:kw,ab,ta,ti,tn,mn,jt,pt OR foot:kw,ab,ta,ti,tn,mn,jt,pt OR triceps:kw,ab,ta,ti,tn,mn,jt,pt OR biceps:kw,ab,ta,ti,tn,mn,jt,pt OR tibialis:kw,ab,ta,ti,tn,mn,jt,pt

3 1 and 2

##### Scopus (1473)

TITLE-ABS-KEY ( ( myasthenia ) AND ( distal OR arm OR wrist OR hand OR finger OR thumb OR leg OR ankle OR foot OR triceps OR biceps OR tibialis ) )

##### Web of Science Core Collection (All Editions) (593)

myasthenia AND (distal or arm or wrist or hand or finger or thumb or leg or ankle or foot or triceps or biceps or tibialis) (All Fields)

#### Publications Included in the Scoping Review

| No. | Year | Reference | Title |
|---|---|---|---|
| 1 | 1997 | Uncini et al. | Distal myasthenia gravis and sensory neuronopathy with anti-50 kDa antibody mimicking sensory-motor neuropathy |
| 2 | 1998 | Janssen et al. | Myasthenic hand |
| 3 | 1999 | Nations et al. | Distal myasthenia gravis |
| 4 | 2000 | Gilad et al. | Bilateral Foot Drop as a Manifestation of Myasthenia Gravis |
| 5 | 2001 | Musser et al. | Distal myasthenia gravis with a decrement, an increment, and denervation. |
| 6 | 2002 | Karacostas et al. | Isolated distal hand weakness as the only presenting symptom of myasthenia gravis |
| 7 | 2003 | Scola et al. | Miastenia grave distal: relato de caso |
| 8 | 2006 | De Carvalho et al. | Longstanding right-hand weakness in a patient with myasthenia gravis |
| 9 | 2007 | Samuraki et al. | Myasthenia gravis presenting with unusual neurogenic muscle atrophy |
| 10 | 2008 | Renard et al. | Distal myasthenia gravis |
| 11 | 2013 | Cirillo et al. | Clinical reasoning: A 62-year-old man with right wrist drop |
| 12 | 2013 | Pardo et al. | Distal myasthenia simulating radial nerve palsy: A case report |
| 13 | 2015 | Fearon et al. | Distal myasthenia gravis presenting as isolated distal myopathy |
| 14 | 2016 | Andrade et al. | Distal Myasthenia Gravis: An unusual presentation with slowly progressive distal weakness and atrophy |
| 15 | 2016 | Rodolico et al. | Myasthenia Gravis: Unusual Presentations and Diagnostic Pitfalls |
| 16 | 2017 | Abraham et al. | Selective or predominant triceps muscle weakness in African–American patients with myasthenia gravis |
| 17 | 2017 | Sousa et al. | Ophthalmoparesis and unilateral finger flexor muscle weakness in seronegative myasthenia gravis |
| 18 | 2018 | Atmaca et al. | Myasthenia gravis with clinical facilitation: a case series |
| 19 | 2019 | Thilak et al. | Isolated bilateral triceps muscle weakness as a presenting complaint in myasthenia gravis: A review |
| 20 | 2021 | Mansukhani et al. | Distal myasthenia gravis – A missed clinical diagnosis |
| 21 | 2021 | Shalabi et al. | A Unique Case of Unilateral Distal Arm Weakness as a Presentation of Myasthenia Gravis |
| 22 | 2022 | Dragusin et al. | Low Fluctuation of Symptoms May Delay Diagnosis of Myasthenia Gravis: A Case Series |
| 23 | 2022 | Alanazy et al. | Finger Flexor Weakness in Myasthenia Gravis |
